# Early Assessment and Intervention for Families Experiencing Homelessness: A randomized trial comparing two parenting programs

**DOI:** 10.1101/2020.12.07.20245563

**Authors:** Paulo A. Graziano, Jamie A. Spiegel, Emily Arcia, Sundari Foundation

## Abstract

**Objective:** As part of a larger community-based, service driven research project, the purpose of this study was to 1) document the clinical needs of sheltered children (i.e., externalizing behavior problems [EBP], trauma symptoms, and developmental status) and 2) examine the effectiveness of time-limited adaptations of Parent Child Interaction Therapy (PCIT) and Child Parent Psychotherapy (CPP).

**Method:** One hundred and forty-four young children (*M*_*age*_ = 3.48, *SD* = 1.09; 56.9% male; 78.5% Black/African American) and their mothers were recruited from a women’s homeless shelter and randomly assigned to a 12-week course of either PCIT or CPP delivered by shelter clinicians on site. Families completed pre and post intervention assessments including mother-reported measures of child EBP and trauma symptoms, and parenting stress. Observational data on maternal verbalizations during a child-led play session were also collected.

**Results:** High rates of clinically elevated EBP (36%), trauma symptoms (47%), and developmental delays (35%) were found. Time-limited PCIT resulted in greater reductions in maternal negative verbalizations and parenting stress, and greater increases in maternal positive verbalizations relative to time-limited CPP. As it relates to child outcomes, both time-limited PCIT and CPP resulted in similar decreases in children’s post-traumatic stress symptoms; however, only time-limited PCIT resulted in significant improvements in EBP.

**Conclusions:** Young sheltered children evidenced high levels of EBP, trauma symptoms and developmental delays. Both time-limited PCIT and CPP offered effective trauma interventions. However, PCIT offered a more comprehensive intervention by more effectively targeting EBP and parenting. Implications of these findings for clinical practice are discussed.

---

Homelessness, defined as a condition in which individuals lack “fixed, regular, and adequate nighttime residence” (McKinney-Vento Act, 2006), is a global problem impacting over 100 million people worldwide (UN-Habitat, 2005). Most troubling, the most recent census data (from 2013) indicates that one in every 30 children in the U.S., or 2.5 million, had experienced homelessness each year (Bassuk et al., 2014). Despite the magnitude of childhood homelessness, there is a dearth of recent empirically-based research assessing the special needs of homeless children and effective supportive interventions to address those needs in shelter environments. Studies most often cited, now more than two decades old, find disproportionally higher rates of unmet health needs (e.g., acute health problems, trauma-related injuries) in children experiencing homelessness than in the general population. Up to 78% of children experiencing homelessness suffer from at least one mental health issue (e.g., depression, behavior problems) along with academic and/or developmental delays (Committee on Community Health Services, 1996; Weinreb et al. 1998). Providing extended mental health services presents unique challenges for children and families experiencing homelessness. Specifically, although shelter stays have lengthened for many families, the most recent scientific study indicates that approximately three quarters of all families experiencing homelessness are “temporary” shelter users (i.e., shelter stays tend to be no more than three months in length; Culhane et al., 2007), meaning that they only have physical access to any given shelter’s services for short periods of time. Moreover, given that over half of all homeless children in the U.S. are under the age of 6 (Samuels et al., 2010), it is particularly important to investigate the feasibility and effectiveness of delivering evidence-based parenting programs within a shelter setting.

## Mental Health Needs of Young Children Experiencing Homelessness

Externalizing behavior problems (EBP), including aggression, defiance, inattention, hyperactivity, and impulsivity are the most common reasons for early childhood mental health referral (Cormier, 2008). In addition to having a highly stable and persistent course starting as early as age 2 (Lee et al., 2008), early-onset EBP are associated with a developmental trajectory of psychosocial impairment, including increased risk for later antisocial behavior (Moffitt et al., 2002), substance use disorders (Lee, et al., 2011), peer rejection (Hoza, 2007), and negative academic outcomes (Loe & Feldman, 2007). Therefore, young children with EBP represent an optimal at-risk population for early intervention.

Children experiencing homelessness are at a higher risk for developing early-onset EBP (Koblinsky et al., 2000) and more severe EBP (Bassuk et al., 1997) than their non-homeless peers. The National Child Traumatic Stress Network reports that “more than one-fifth of homeless preschoolers have emotional problems serious enough to require professional care, but less than one-third receive any treatment” (Bussuk & Friedman, 2005, p. 2). Notably, such estimates likely represent an underestimation of comparative risk of mental health difficulties, due to reliance on comparing children experiencing homelessness to low income youths as opposed to all age-matched peers (Bassuk et al., 2015).

Homelessness is associated with a higher incidence of exposure to traumatic events (Anooshian, 2005; Cowan, 2007; Guarino & Bassuk, 2010; Hicks-Coolick et al., 2003; Perlman & Fantuzzo, 2010), complex trauma (i.e., polyvictimization or prolonged exposure to trauma), and adverse childhood experiences, including poverty, family and housing instability, separation from caregivers, community violence, and decreased access to health care and educational services (Masten et al., 1997; Panter-Brick, 2004; Shelton et al., 2015; Zlotnick, 2009).

Indeed, 20% of youths experience some form of trauma and approximately half of trauma survivors experience polyvictimization (e.g., Saunders & Adams, 2014)^1^. The varied presentation of post-traumatic responses is highlighted by the extensive number of possible combinations of symptoms of post-traumatic stress disorder delineated in the DSM 5 (American Psychiatric Association, 2013) which include EBP as well as internalizing symptoms. Although experiences vary, homelessness represents a complex stressor to which the majority of children respond at a minimum with worries about the safety of themselves and their families (National Center on Family Homelessness, 1999). Given the multitude of serious difficulties associated with childhood homelessness, it is imperative to test interventions that might successfully target trauma symptoms. As such, parent-based early intervention programs with their proven efficacy in the general population (Eyberg et al., 2001), offer a treatment option worth investigating.

## Parenting Challenges

As with children, parents vary in their response to homelessness. Whereas some parents demonstrate resiliency and positive parenting practices other parents struggle, or their pre-existing parenting difficulties are exacerbated in the face of the increased challenges imposed by homelessness. Overall, studies suggest that homelessness is associated with increased parental frustration and decreased confidence in parenting (Lee et al., 2010), decreased parental warmth, decreased positive parent-child interactions (Koblinsky et al., 1997), increased incidence of negative parenting behaviors including violence or aggression (Lindsey, 1998; Torquati, 2002), and a consequent increased involvement with child protective services and foster care placement (McChesney, 1995; Fantuzzo & Perlman, 2007).

Parent-child relationships can be influenced by parents’ own chronic medical, mental health, and substance abuse difficulties that are exacerbated when experiencing homelessness (Arangua et al., 2005; Caton et al., 2005; Lee et al., 2010; Shinn & Weitzman, 1996; Weinreb et al., 2006). Homeless families are also more likely than their homed counterparts to be headed by single mothers who often have received minimal education and job training (Bassuk et al., 1997; Burt et al., 1997), and who have often experienced negative parenting role-models, substantial childhood trauma, and/or recent domestic violence (e.g., Anooshian, 2005; Swick & Williams, 2010). These parental risk factors combine to create unduly difficult circumstances for the development of positive parenting practices, which are further compromised by the fact that children experiencing homelessness often present with numerous developmental, educational, social, emotional, and EBP difficulties. Finally, the environmental constraints of the shelter itself may exacerbate parenting difficulties. Parents experiencing homelessness often report feeling judged by other residents and shelter staff for their parenting practices (Lindsey 1998). For parents who have relied on corporal punishment as a disciplinary strategy, feelings of frustration and lack of control when living in a shelter can be exacerbated by the fact that shelters typically impose both child-level behavioral expectations and restrictions on the use of corporal punishment (Lindsey, 1998; Swick & Williams, 2010). The numerous risk factors faced by children and parents experiencing homelessness coupled with the influence that parent-child relationships have on children’s well-being highlights the importance of promoting positive parenting strategies in shelter environments.

## Evidence-based Parenting Programs

Behavioral parent training (BPT) programs are among the most well-established evidence-based interventions for EBP in young children (Eyberg et al., 2008). BPT programs reduce EBP by promoting positive parent-child interactions and parental consistency in the use of non-corporal disciplinary strategies such as time outs. Large effect sizes on both behavioral outcomes (Kaminski et al., 2008) and trauma symptoms (e.g., Pearl et al., 2012) have been documented across various BPT programs. One such evidence-based BPT program, and the focus of the current study, is Parent-Child Interaction Therapy (PCIT; Eyberg et al., 2001). PCIT is divided into two phases: child directed interaction and parent directed interaction (see McNeil & Hembree-Kigin, 2010 for a comprehensive description of the skills taught in PCIT). Although originally designed to treat EBP, PCIT has been demonstrated to be effective in the treatment of children exposed to a variety of early childhood stressors, including domestic violence (Borrego et al., 2008; Pearl, 2008), caregiver psychopathology (e.g., Babinski et al., 2014; Chengappa et al., 2017; Pemberton et al., 2013), and early childhood maltreatment (e.g., Pearl et al., 2012; Self-Brown et al., 2012). Hence, on the one hand, PCIT may be an ideal intervention for children and their families who are experiencing homelessness. On the other hand, traditional PCIT might be difficult to implement in its totality with sheltered families because treatment averages 20.5 sessions given that shelter stays may not be long enough to satisfy a strict “mastery” criteria (Lieneman et al., 2019).

Child Parent Psychotherapy (CPP; Lieberman et al., 2005) is an evidence-based intervention designed for the treatment of early childhood trauma. The intervention is divided into three phases: assessment and engagement, core intervention, and recapitulation and termination. CPP has been demonstrated to be effective in improving parent-child interactions, children’s cognitive functioning (Lieberman et al., 2015), and trauma symptoms (Lieberman et al., 2005). As with PCIT, CPP has been effectively utilized in the presence of several early life stressors, including impoverishment, caregiver psychopathology (Cicchetti et al., 2000), comorbid anxiety and depression symptoms, and placement in the foster care system (Lieberman et al., 2015). However, to date, CPP has not been examined within the context of homelessness; likely because full implementation can require 50 to 52 weeks and homeless families may be unable to stay in a given shelter for more than a few months (Culhane et al., 2011).

The viability of shortening the delivery of evidence-based parenting programs to maximize rapid improvement and cost-effectiveness has received increased attention in the recent literature (Hare & Graziano, *in press*; Mersky et al., 2015) and such a time-limited approach to PCIT and CPP might be particularly well suited to families experiencing homelessness. Time-limited PCIT entails a standard number of sessions that do not require that caregivers meet “mastery” criteria prior to graduation. In effect, time-limited PCIT of 10 to 12 sessions has demonstrated promising results both in improving parent-child interactions and child compliance, thereby diminishing EBP (Graziano et al., 2020; Nixon et al., 2003; Thomas & Zimmer-Gembeck, 2012). A time-limited adaptation of CPP would maintain the requirement that families pass through all three phases of intervention in a standard, abbreviated, number of sessions. To our knowledge no study to date has examined the effectiveness of time-limited CPP and neither CPP nor PCIT have been examined with sheltered families. Given the effectiveness of both PCIT and CPP with at-risk populations, and the clear applicability of time-limited versions of such programs, an empirical investigation of their feasibility and effectiveness with homeless families was warranted.

## Goals of the Current Study

Taken together, it is clear that children and families experiencing homelessness possess a wide range of needs, compounded by stressors leading up to and including homelessness, that negatively affect the wellbeing of children, parenting and the parent-child relationship. Given that children under 6 years of age represent the largest segment of children experiencing homelessness, it is particularly important to provide early needs assessments and evaluate evidence-based parenting interventions that might be appropriate for this vulnerable population. Thus, as part of a larger community-based service driven research project (Arcia, 2020), the current study sought to 1) document the clinical needs of sheltered homeless children (i.e., EBP, trauma symptoms, developmental status), and 2) examine the promise of two established parenting programs to support sheltered children and mothers experiencing homelessness. Following a clinical assessment, families were randomized to receive 12 sessions of either 1) PCIT or 2) CPP delivered within the homeless shelter. First, we hypothesized high rates of clinically elevated levels of EBP, trauma symptoms, and developmental delays in this at-risk population. Second, as it relates to the intervention, we hypothesized that both time-limited programs would be feasible to implement, be well attended, and receive high consumer/intervention satisfaction scores. Given CPP’s focus on trauma, we expected children randomized to time-limited CPP to have greater reductions in parent reported trauma symptoms relative to time-limited PCIT. On the other hand, given PCIT’s live coaching framework both as it relates to improving the child-parent relationship and discipline, we expected parents in time-limited PCIT to experience greater gains in positive parenting skills, reducing negative parenting, and decreasing child EBP relative to time-limited CPP. Lastly, given the supportive nature of PCIT and CPP, we expected parents from both interventions to experience similar reductions in parenting stress.

## Method

### Participants and Recruitment

The current study, which was part of a larger service driven, community based, research project, took place at (masked for review), the largest women’s shelter in the State of (masked for review). At the time of implementation, the women and children sheltered at (masked for review) were predominately Black (66% - Black/Non-Hispanic, 22% White Hispanic, 6% Black/Hispanic, 5% White Non-Hispanic, and 1% other ethnicities). Ninety percent of the women at the shelter were victims of adult violence (e.g., domestic violence) and/or childhood violence including neglect (40%), physical abuse (45%), and sexual abuse (50%). To qualify for the current study, families were required to (a) have a child between the ages of 18-months and 5-years-of-age and (b) have a mother who spoke English or Spanish. Though mothers could elect to receive clinical services without participating in the service driven research, virtually all mothers entering the shelter provided written consent for the results of their initial screening assessment and response to intervention be used in research (*n* = 1,025). Exclusionary criteria, for the current randomized study, included children a) not being in the target age range, b) already receiving therapy services elsewhere, c) having a sibling currently receiving another intervention at the shelter, or d) requiring referral for other services (e.g., applied behavior analysis due to suspected Autism Spectrum Disorder). Accordingly, of the 1,025 children that were screened in the shelter, 877 were excluded from this randomized study (see Figure 1 for a consort diagram outlining study enrollment). It is important to note that all children who were excluded from this randomized trial were offered age appropriate clinical services, based on their initial assessment. For example, PCIT was offered for children ages 6 to 7 with elevated EBP, Trauma Focused Cognitive Behavioral Therapy was offered for children over the age 7, and referrals were made as appropriate to third party providers. See Arcia (2020) for details regarding the additional therapeutic services, clinical needs, and outcome data for non-randomized sample.

**Figure 1.**
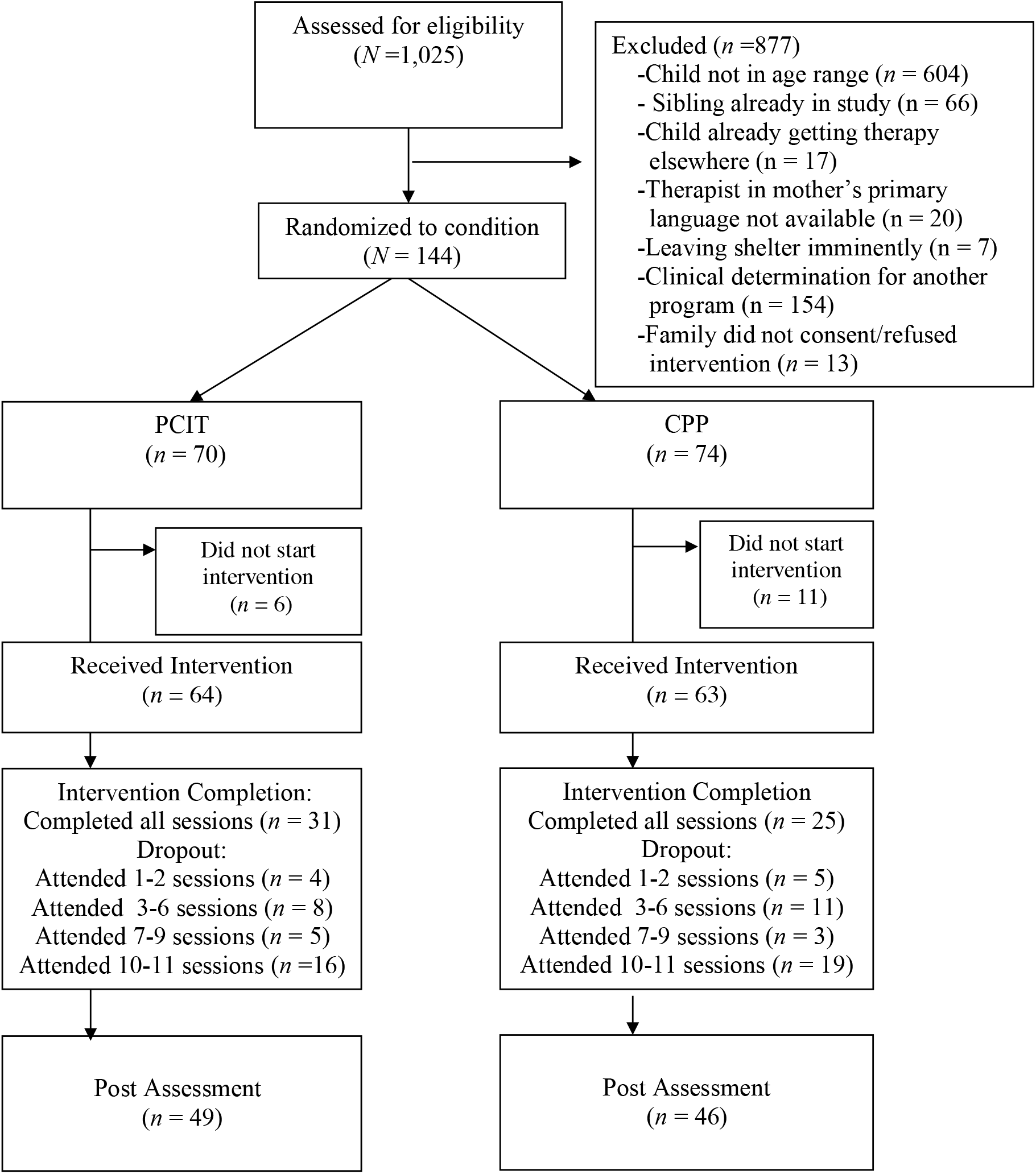
Consort Flow Diagram.

The participating sample consisted of 144 young children whose mothers provided consent to participate in the study. Children had a mean age of 3.48 years (range: 18 months to 5.75 years of age, *SD* = 1.09 years), and most were boys (56.9%) and Black/African American (78.5%). Only one child was currently or had ever taken psychotropic medication. See Table 1 for other descriptive sample data.

**Table 1.** Participant Baseline Demographic Variables by Initial Intervention Assignment.

|  | Total Sample<br>(N = 144) | PCIT<br>(n = 70) | CPP<br>(n = 74) |
| --- | --- | --- | --- |
| **Demographic Variables** |  |  |  |
| Child sex (% male)* | 56.9 | 64.3 | 50 |
| Mean Child age (SD) | 3.48 (1.09) | 3.34 (1.10) | 3.61 (1.08) |
| Child Race (%) |  |  |  |
| Black | 78.5 | 77.1 | 79.7 |
| White | 20.1 | 22.9 | 17.6 |
| Biracial | 1.4 | 0.0 | 2.7 |
| Child Ethnicity (%) |  |  |  |
| Latinx | 27.1 | 30 | 24.3 |
| Non-Latinx | 72.9 | 70 | 75.7 |
| Maternal Education (%) |  |  |  |
| Some High School | 45.1 | 48.6 | 41.9 |
| High School Diploma/GED | 38.2 | 35.7 | 40.5 |
| Technical Degree | 4.9 | 1.4 | 8.1 |
| Some College | 9.0 | 14.3 | 4.1 |
| Associate's Degree | 0.7 | 0 | 1.4 |
| Bachelor's Degree | 2.1 | 0 | 4.1 |
| Maternal Employment Status (%)* |  |  |  |
| Unemployed | 71.5 | 61.6 | 82.2 |
| Employed | 28.5 | 38.4 | 17.8 |
| Home language (%) |  |  |  |
| Monolingual (English only) | 81.3 | 79.3 | 83.1 |
| Monolingual (Spanish only) | 3.3 | 3.4 | 3.1 |
| Monolingual (Other Language) | 0.8 | 0 | 1.5 |
| Bilingual (Spanish/English) | 13.8 | 17.2 | 10.8 |
| Bilingual (English/Other Language) | 0.8 | 0 | 1.5 |
| Department of Child and Families Involvement (%) |  |  |  |
| Present | 16 | 18.6 | 13.5 |
| Past | 50 | 48.8 | 51.2 |

*Note.* Values enclosed in parentheses represent standard deviations. \*chi-square analyses indicated a significant difference among the intervention groups on maternal employment status, $\chi^2(1, 144) = 11.23, p < .01$ and marginally on child sex, $\chi^2(1, 144) = 2.99, p = .08$ . PCIT = Parent-Child Interaction Therapy, CPP = Child Parent Psychotherapy.

### Study Design and Procedure

This study was approved by the University’s Institutional Review Board. Participants were randomized to time limited PCIT (*n* = 70) or time-limited CPP (*n* = 74) without stratification using a randomly generated number table following their pre-intervention assessment. Clinicians at (masked for review) who delivered the interventions, in the mother’s preferred language, were master’s level licensed clinical staff or therapists in training who were certified or in the process of receiving their certification in PCIT or CPP. For PCIT, counselors received weekly supervision by a licensed clinical psychologist, who was a certified trainer by PCIT International. For CPP, a licensed mental health counselor who had completed CPP training provided biweekly supervision and biweekly consultation calls with a national leading CPP trainer. Additionally, it is important to note that consistent with what is common in community trials, a portion of cases (35%) were seen by a therapist cross-trained in both CPP and PCIT. Due to the setting, it was not possible to record sessions to systematically measure intervention fidelity and/or intervention contamination. However, clinicians for each intervention modality completed content checklists for each session and all analyses were co-varied for therapists’ cross-training status.

At intake clinicians and trained staff administered an assessment protocol that lasted approximately two hours and included: a) a biopsychosocial interview of mothers that gathered relevant background information on the family, b) questionnaires on children’s EBP, trauma experiences and symptoms, c) questionnaires on maternal parenting stress, and d) videotaped observations of three 5-minute standard parent-child interaction situations that varied in the degree of parental control expected (child-led play, parent-led play, and clean-up). During the same visit, a clinician administered the Battelle Developmental Inventory, 2^nd^ Edition (BDI-2; Newborg, 2005), a comprehensive assessment tool used to assess developmental skills in children aged birth to 7-years 11-months. With the exception of the BDI-2, which was not repeated, families completed a similar post-intervention assessment upon completion of intervention (i.e., 12 sessions) or four months after the start of the intervention (mean time to complete post-intervention assessment = 4.77 months, *SD* = 1.92 months). Families were given small incentives such as a small toy to the child, or a small gift to the parent upon completion of the assessments, and all interventions were provided at no cost.

Comparison of the demographic characteristics of the two groups indicated significant differences between the groups on only two variables which were later co-varied on all analyses. As seen in Table 1, unemployed mothers were more likely to be randomized to time limited CPP and boys were marginally more likely to be randomized to time limited PCIT.

### Intervention Description and Adaptation

#### Parent-child Interaction therapy

(PCIT; Eyberg & Robinson, 1982). PCIT is a manualized evidence-based BPT program that integrates social learning and attachment theories. In PCIT, parents proceed through two distinct phases: Child-Directed Interaction (CDI), which resembles traditional play therapy, and Parent-Directed Interaction (PDI), which resembles clinical behavior therapy. During CDI, parents follow their child’s lead in play by using the non-directive PRIDE (i.e., *do skills*): <u>P</u>raising the child, <u>R</u>eflecting the child’s statements, Imitating the child’s play, <u>D</u>escribing the child’s behavior, and using <u>E</u>njoyment. Parents learn to apply PRIDE skills to the child’s appropriate play and ignore undesirable behaviors, and are taught to avoid verbalizations that take the lead away from the child during the play (i.e., *don’t skills*), including questions, commands, and negative statements (e.g., criticism). During PDI, parents set limits to reduce child noncompliance and negative behavior. They learn to use effective commands and consistently follow through with timeout for noncompliance. During all sessions, the therapist coaches each parent *in vivo* in their use of the CDI and PDI skills with their child. Of note, in the current study therapists coached parents in the same room given that the homeless shelter was not equipped with one-way mirror rooms that traditionally have been used in PCIT. In traditional PCIT, parents must also meet “mastery” criteria after each phase to progress and complete treatment. Mastery of CDI is met when parents are able to demonstrate a high level of positive parenting skills during a five-minute observation period. Mastery of PDI consists of limiting negative parenting and successfully implementing appropriate consequences during another five-minute interaction with their child. Consequently, treatment course can vary greatly in length with the largest PCIT study (*n* = 1,318), to our knowledge, averaging 20.5 weekly sessions (Lieneman et al., 2019). For a full detailed description of traditional PCIT see Zisser & Eyberg (2010). The only adaptations the current study made, similar to prior work (Thomas & Zimmer-Gembeck, 2012), was to limit the number of sessions to 12 and to not require that mothers meet “mastery” criteria to progress and complete treatment. Thus, all families randomized to time-limited PCIT received 12 total weekly sessions (6 sessions of CDI and 6 sessions of PDI).

#### Child-parent psychotherapy

(CPP; Lieberman et al., 2005). CPP is a relationship-based treatment that was originally developed to improve the psychological and relational functioning of young children exposed to trauma. CPP integrates attachment, cognitive-behavioral, social-learning, and psychodynamic theories and focuses on the child-parent relationship as a way to improve the child’s adaptive functioning. Various intervention strategies are flexibly employed in CPP including a) joint construction of a trauma narrative, use of play and language to identify and address traumatic triggers, and building of an emotional vocabulary; b) unstructured, supportive developmental guidance to provide psychoeducation regarding children’s safety and developmental needs, c) modeling protective behavior, d) insight-oriented interpretations to increase self-understanding in parent and child e) emotional support and affect regulation, and f) assistance with daily living issues, including crisis intervention, case management, and service referrals. CPP is conducted with the parent-child dyad in unstructured weekly hour-long sessions which allows therapists to flexibly tailor each session to the needs of the individual family. CPP was originally designed as a yearlong intervention in which therapists move through three phases: assessment and engagement, core intervention, and recapitulation and termination. See Lakatos et al. (2019) for a full description of each of the phases of CPP. Although the intention of CPP is for the parent-child dyad to complete 50 sessions, the average number of sessions completed actually tends to be much lower at about 21 sessions (Hagan et al., 2017), which is similar to PCIT. The only adaptions to CPP made in the current study were to a) limit the number of sessions to 12 (to equate the intervention dose to that of PCIT) and b) make sure that therapists progressed families across all phases of CPP prior to termination. The flexibility of CPP was maintained in terms of no imposed number of sessions per phase.

### Measures of Feasibility and Acceptability

#### Intervention fidelity

For both modalities therapists completed content checklists for each session. Intervention supervisors randomly checked 20% of those sessions by comparing the electronic medical records (ERM) intervention session notes to the checklists. Weekly or biweekly (if consultation calls were occurring) group supervision lasting between 1-2 hours was provided by an expert in each respective modality.

#### Intervention Completion and Attendance

Attendance for each session was measured from therapists’ contact notes within the EMR system. Intervention completion rates were calculated based on the percentage of families that completed 12 sessions within a 16-week period. The current study also calculated the percentage of families that eventually completed almost all intervention beyond the 16-week assessment period defined as completing at least 10 out of the 12 sessions.

#### Consumer/intervention satisfaction

Parents provided ratings of satisfaction at post-intervention by completing selected items from the Therapy Attitude Inventory (Brestan et al., 1999). Parents indicated their degree of satisfaction across a five-point Likert scale regarding a) improvements in the parent-child relationship b) progress the child has made in his/her general behavior, c) progress the child has made in his/her trauma symptoms or traumatic/stressful experiences, d) general feeling about the program parent participated in, and e) how likely the parent was to recommend the program to others. The mean level of satisfaction was calculated across these five items (α = .72).

### Parenting Outcomes

#### Parenting Stress

Mothers completed the Parenting Stress Index-Short Form (PSI-SF; Abidin, 1983). The PSI-SF is a widely used 36-item self-report instrument for parents of children ages 1 month to 12 years measuring parental stress (Abidin, 1983). The PSI-SF total raw score was used to measure overall parenting stress (α’s for the current study = .85-.90).

#### Parenting Skills

The *Dyadic Parent-Child Interaction Coding System-4*^*th*^ *Edition* (DPICS-IV; Eyberg et al., 2013), an established behavioral coding system was used to measure the quality of parent-child interactions during a 5-minute child-led play session. Consistent with prior research, we coded and created a composite of positive parenting verbalizations (behavior descriptions, reflections, praises) and negative parenting verbalizations (questions, commands, and negative talk) used during child-led play. To account for mothers’ total verbalizations, including neutral verbalizations, the current study also used a proportion score ranging from 0 to 1 for both positive and negative verbalizations (e.g., the total number of positive verbalizations was divided by the total number of positive, negative, and neutral verbalizations; Bagner et al., 2016). Staff coders, who were masked to intervention status, were trained to 80% agreement with a criterion tape and 20% of the observations were coded a second time. Reliability for the *positive* and *negative verbalizations* were excellent *(r’*s range from .96 to .97).

### Child Outcomes

#### Externalizing Behavior Problems (EBP)

Mothers completed the *Eyberg Child Behavior Inventory* (ECBI; Eyberg & Ross, 1978), a 36-item questionnaire that is designed to assess the presence of externalizing problems in children ages 2 through 16 years. The total intensity scale raw score was used in the current study as the main measure of EBP (α’s = .84-.93).

#### Post-Traumatic Stress Symptoms (PTSS)

Mothers of children ages 3 and older completed the *Child and Adolescent Trauma Screen-Caregiver (CATS-C;* Sachser et al., 2017), which consists of an event checklist of 15 potentially traumatic events, as well as the frequency of each of the 20 PTSS, based on DSM-5 criteria (American Psychiatric Association, 2013). Responses are provided based on a 4-point Likert-scale ranging from 0 = ‘never’ to 3 = ‘almost always’ with higher scores indicative of greater PTSS. The total severity score of PTSS was used in the current study (α’s = .72-.75).

### Data Analytic Plan

All analyses were conducted using Statistical Package for the Social Sciences, version 20 (SPSS 26). There was only 5% missing data for pre-intervention variables. Approximately 36% of post-intervention data were missing due to families who dropped out of intervention and did not complete any post-intervention assessments. Families with completed versus partial data did not differ on any demographic variables. Little’s Missing Completely at Random (MCAR) test confirmed that the data were not missing at random, χ^2^(27) = 57.50, *p* < .01. Therefore, as recommended in clinical trials, intent to treat analyses with the use of multiple imputation was used (Little & Yau, 1996; Rubin, 1988; Von Hippel, 2020). Initial analyses focused on describing the special needs of this population in terms of providing the percentage of children who were clinically elevated in terms of EBP, trauma symptoms, and experiencing developmental delays. Next, we examined differences in intervention fidelity, completion, attendance, and intervention satisfaction between the intervention groups via chi-square analyses or ANCOVAs. For the primary analyses, multiple repeated measures ANCOVAs were conducted to compare families who were randomized to PCIT versus CPP in terms of parenting and child outcomes. Bonferroni corrections to minimize Type 1 error were utilized while Cohen’s *d* effect size (ES) estimates for within-subjects were calculated separately for each intervention by comparing baseline to post-intervention.

## Results

### Special Needs Profile at Pre-Intervention Assessment

In terms of EBP, 36.3% of the children had total raw scores in the clinical range as reported by mothers on the ECBI (i.e., score of 131 or higher). In terms of trauma symptoms, 47.3% of the children had total raw scores in the clinical range as reported by mothers on the CATS (i.e., score of 12 or higher on PTSD severity). As it relates to children’s developmental status, 35.7% of children had overall scores on the BDI-2 indicating a suspected developmental delay requiring a referral for further evaluation Referral rates for the adaptive, personal-social, communication, motor, and cognitive domains of the BDI-2 were 35.4%, 40.3%, 33.3%, 9%, and 32.6%, respectively. Of note, 44.2% of children were clinically elevated in at least one of these domains with 16.7% being elevated across all three domains measured (ECBI, CATS, BDI-2).

### Intervention Fidelity, Completion, Attendance, and Satisfaction

Overall intervention fidelity of the content covered across PCIT sessions was high (*M* = 96%; range 82-100%). Procedural and content fidelity of CPP were also high (procedural fidelity *M* = 92%; range 75-100% and content fidelity *M* = 93%; range 79-100%). In terms of intervention completion, it is important to note that 13.7% of families in time-limited CPP and 8.5% of families in time-limited PCIT dropped out after randomization and never initiated any intervention (see Figure 1). Of families that initiated intervention, 48.4% of families in time-limited PCIT (*n* = 31) completed the intervention (i.e., 12 sessions) within 16 weeks compared to 39.7% of families in time-limited CPP (*n* = 25). Completion rates did not differ significantly between the time-limited PCIT and time-limited CPP group (χ^2^ = 1.01, *p* > .05). Of note, 71.4% of families in time-limited CPP and 76.6% of families in time-limited PCIT eventually completed the intervention (i.e., after 16 weeks and at least 10 out of the 12 sessions completed). The average number of intervention sessions attended between groups also did not differ (PCIT = 9.62 sessions, *SE* = .48, and CPP = 9.23 sessions, *SE* = .48). Lastly, parents reported high levels of satisfaction across both time-limited PCIT (*M* = 4.24 *SE* = .11) and time-limited CPP (*M* = 4.25, *SD* = .12).

### Parenting Outcomes

As indicated in Table 2 and graphically presented in Figure 2, whereby time indicates pre- and post-intervention assessments, a significant time by group interaction was noted for total negative parenting verbalizations, F (1, 139) = 7.80, *p* <.01, partial eta-squared = .05, total positive parenting verbalizations, F (1, 139) = 34.80, *p* <.001, partial eta-squared = .20, proportion of negative, F (1, 139) = 42.24, *p* <.001, partial eta-squared = .23, and positive parenting verbalizations, F (1, 139) = 50.31, *p* <.001, partial eta-squared = .27. Specifically, mothers in time-limited PCIT had significantly greater reductions in total proportion of negative verbalizations as well as increases in total and proportion of positive verbalizations compared to mothers in time-limited CPP at post-intervention assessment from pre-intervention levels. As seen by the effect sizes in Table 2, it is important to note that mothers in time-limited CPP experienced significant improvements in total and proportion of positive parenting verbalizations (Cohen’s *d* = .50 and .61, respectively) but at a lower level relative to the large gains seen in mothers in time-limited PCIT (*d* = 1.72 and 2.14, respectively). It is also important to note that while mothers in time-limited CPP experienced some reductions in proportion of negative parenting verbalizations, their reduction in total negative parenting verbalizations was not significant as the 95% confidence interval for the effect size contained zero.

**Table 2.** Results of ANCOVA Analyses Examining Parent and Child Outcomes of Time-limited PCIT and CPP Controlling for Sex and Maternal Employment Status

|  | Pre-Intervention<br>*M (SE)* | Post-Intervention<br>*M (SE)* | Time<br>Effect<br>*F* | Time x<br>Group<br>*F* | Pre-Post<br>*d* [95% CI] |
| --- | --- | --- | --- | --- | --- |
| **Parenting Outcomes** |  |  |  |  |  |
| Parenting Stress: PSI total stress raw score (P) |  | --- | **50.85\*\*\*** | **4.06\*** | --- |
| PCIT | 91.96 (2.63) | 74.16 (2.06) | --- | --- | -.90 [-.55, -1.24] |
| CPP | 86.66 (2.56) | 75.50 (2.00) | --- | --- | -.56 [-.23, -.89] |
| Proportion of Negative Parenting Verbalizations (O) |  | --- | **40.85\*\*\*** | **18.70\*\*\*** | --- |
| PCIT | .55 (.02) | .27 (.02) | --- | --- | -1.67 [-1.28, -2.05] |
| CPP | .57 (.02) | .51 (.02) | --- | --- | -.38 [-.05, -.70] |
| Proportion of Positive Parenting Verbalizations (O) |  | --- | **64.92\*\*\*** | **50.31\*\*\*** | --- |
| PCIT | .10 (.01) | .39 (.02) | --- | --- | 2.15 [1.73, 2.56] |
| CPP | .10 (.01) | .18 (.02) | --- | --- | .61 [.28, .94] |
| Total Negative Parenting Verbalizations (O) |  |  | 1.90 | **7.80\*\*** |  |
| PCIT | 29.86 (2.07) | 18.28 (1.91) | --- | --- | -.70 [-.35, -1.03] |
| CPP | 34.98 (2.01) | 31.54 (1.86) | --- | --- | -.21 [.12, .53] |
| Total Positive Parenting Verbalizations (O) |  |  | **60.91\*\*\*** | **34.80\*\*\*** |  |
| PCIT | 5.65 (.69) | 20.26 (1.26) | --- | --- | 1.72 [1.32, 2.09] |
| CPP | 6.14 (.67) | 10.43 (1.22) | --- | --- | .50 [.17, .83] |
| **Child Outcomes** |  |  |  |  |  |
| Externalizing Behavior Problems: ECBI total raw score (P) |  | --- | **6.34\*** | **5.66\*** | --- |
| PCIT | 116.60 (5.01) | 102.12 (4.42) | --- | --- | -.37 [-.03, -.70] |
| CPP | 116.23 (4.86) | 115.19 (4.30) | --- | --- | -.02 [.30, -.35] |
| Post-traumatic stress symptoms: total severity score (P) |  | --- | **12.73\*\*** | .11 | --- |
| PCIT | 11.72 (1.19) | 7.92 (.78) | --- | --- | -.45 [-.11, -.78] |
| CPP | 10.67 (1.10) | 7.38 (.73) | --- | --- | -.41 [-.08, -.73] |

*Note.* Means and SEs are marginal estimates after controlling for sex, maternal employment status, and cross-training therapist status. P = Parent report, O = Observation, PCIT = Parent-Child Interaction Therapy, CPP = Child Parent Psychotherapy. Cohen's standardized *d* for group x time effect is for each intervention group. \**p*<.05, \*\**p*<.01, \*\*\**p*<.001.

**Figure 2.**
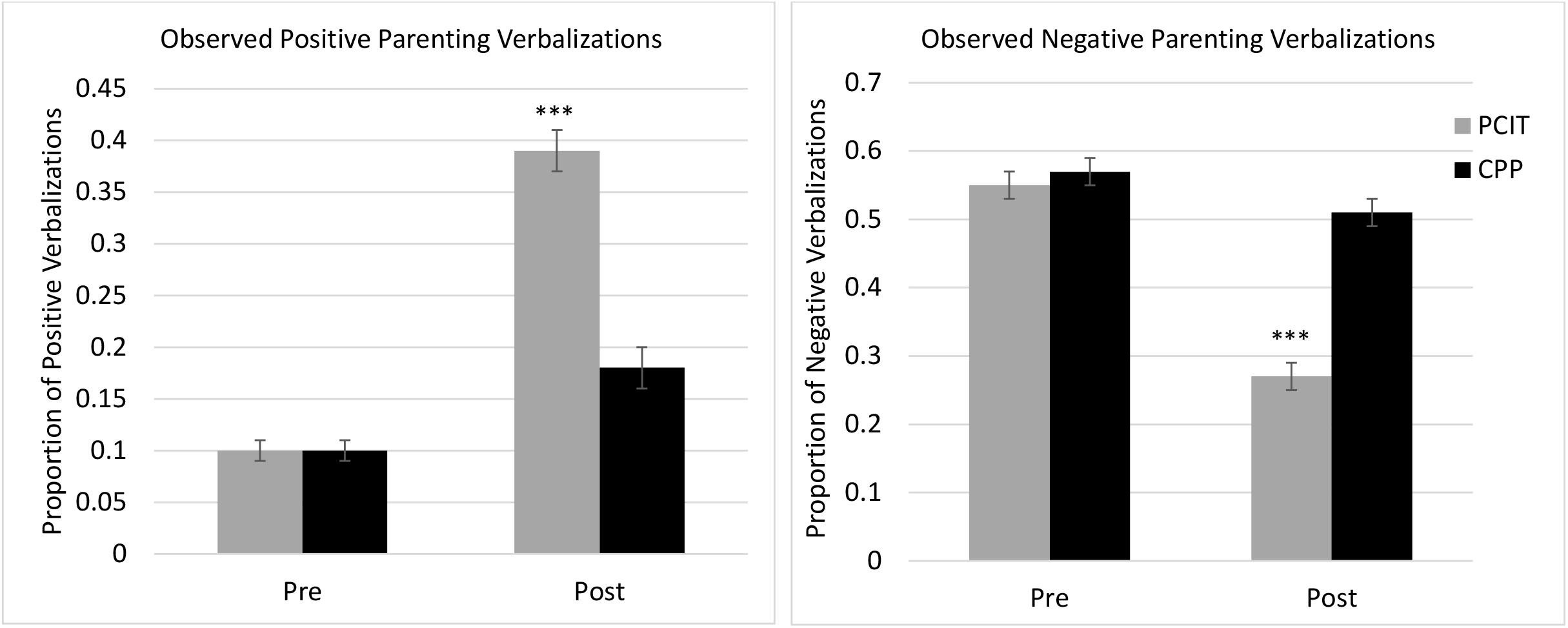
Changes in observed positive and negative parenting verbalizations during child-led play session. ***indicates a significant intervention by group effect at *p* <.001.

**Figure 3.**
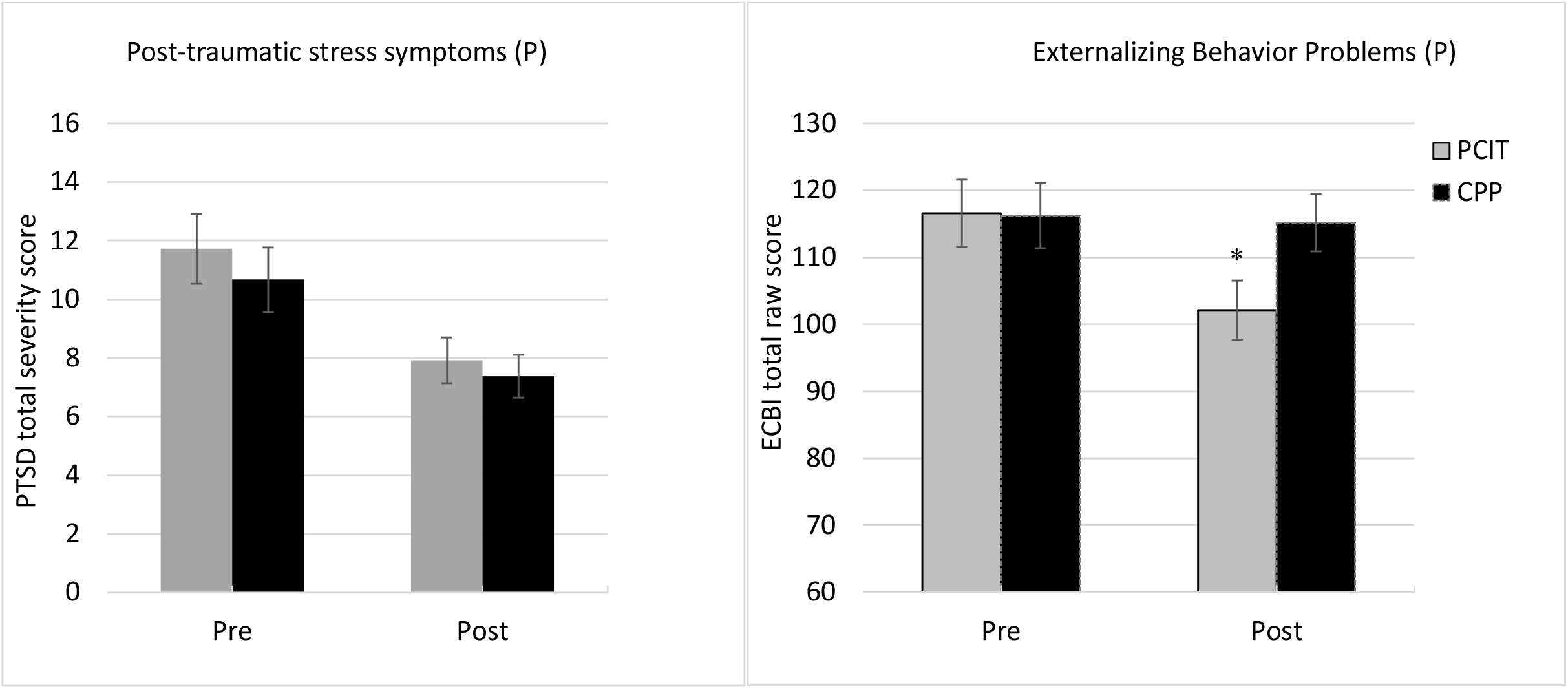
Changes in children’s post-traumatic stress symptoms and behavior problems as reported by parents. *indicates a significant intervention by group effect at *p* <.05.

A significant time by group interaction was also found for parenting stress, F (1, 139) = 4.06, *p* < .05, partial eta-squared = .03. In other words, mothers in time-limited PCIT reported significantly greater reductions in overall parenting stress relative to mothers in time-limited CPP. An examination of the effect sizes showed that mothers in time-limited CPP experienced significant reductions in parenting stress (*d* = -.56) but at a lower magnitude relative to mothers in time-limited PCIT (*d* = -.90).

### Child Behavior Outcomes

As indicated in Table 2, a significant time by group interaction was noted for overall EBP, F (1, 139) = 5.66, *p* <.05, partial eta-squared = .04, such that mothers in time-limited PCIT reported significantly greater reductions in their children’s EBP (*d* = -.36) compared to mothers in time-limited CPP (*d* = -.03). It is important to note that the 95% confidence interval for the effect size of EBP for mothers in time-limited CPP included zero indicating that the reduction in children’s EBP was not statistically significant. Lastly, as it relates to children’s post-traumatic stress symptoms (only for children ages 3 and older), no time by group effect was found. Rather, an overall time effect was found, F(1, 83) = 12.73, p <.001, partial-eta squared = .13, indicating a decrease in post-traumatic stress symptoms reported by mothers equivalent in magnitude across both time-limited PCIT (*d* = -.45) and time-limited CPP (*d* = -.41).

## Discussion

The current study updates and documents the high level of clinical needs in young children experiencing homelessness and demonstrates the value of providing evidence based supportive interventions for sheltered children and families, particularly where mothers have a history of violence and trauma. It also represents the first randomized trial, to our knowledge, examining the efficacy of Parent-Child Interaction Therapy (PCIT) and Child Parent Psychotherapy (CPP) in a homeless shelter. Further, the use of time-limited versions of PCIT and CPP constitutes the first randomized comparison of two abbreviated versions of well-established early intervention programs in an at-risk population. In regard to needs assessment, it is important to point out the high rates of clinically elevated EBP (36%), trauma symptoms (47%), and developmental delays (35%) found in our sample with 16.7% of the children being clinically elevated across all three domains. As it relates to our intervention, both time-limited PCIT and CPP were successfully implemented within the homeless shelter as evidenced by high fidelity rates as well as high satisfaction ratings by mothers. Completion rates and average attendance were similar across both time-limited PCIT and CPP. As it relates to parenting outcomes, time-limited PCIT resulted in greater reductions in maternal negative verbalizations and parenting stress, and greater increases in maternal positive verbalizations compared to time-limited CPP. As it relates to child outcomes, both time-limited PCIT and CPP resulted in significant decreases in children’s post-traumatic stress symptoms; however, only time-limited PCIT resulted in significant improvements in EBP. These findings are discussed further below.

A significant issue in the field of early intervention has been transportability and access to evidence-based programs to those with the greatest needs (Hershell et al., 2004; Silverman et al., 2004). Families experiencing homelessness arguably represent a population with the greatest needs not just in terms of basic and physical needs (e.g., Arangua et al., 2005), but also as it relates to mental health services (e.g., Bussuk & Friedman, 2005; Lee et al., 2010). There has been a dearth of research on the needs of sheltered children over the past 20 years (Hershell et al., 2004; Silverman et al., 2004). The current study takes a crucial step, not only in documenting the high levels of clinical need in young sheltered children such as elevated rates of EBP, trauma, and developmental delays, but also in showing the feasibility of providing in-house therapist training/supervision to aid in the delivery of two well-established evidence-based interventions within a homeless shelter. Completion rates of time-limited PCIT (75%) and CPP (68%) delivered within the homeless shelter were comparable to if not slightly better than those of previous university- or community-based trials of the same or similar parenting programs which typically document dropout rates ranging from 35% to 50% (Chaffin et al., 2009; Danko et al., 2016; Eyberg et al., 2001; McCabe & Yeh, 2009). Thus, a homeless shelter such as (masked for review) which can provide these parenting services in-house has tremendous advantages in terms of circumventing common barriers to providing interventions to this population, most notably engagement in the face of multiple, complex needs and stressors faced by parents, time limitations, lack of resources of both shelters and those they serve, and transportation. Providing free, in-house assessments and supportive parenting programs eliminated barriers to accessing services and allowed flexiblity in terms of scheduling and rescheduling weekly sessions, which helped address the well documented attendance difficulties of families participating in other parenting programs (Axford et al., 2012; Baker et al., 2011). Finally, the use of time-limited interventions also likely contributed to the reduced dropout rates, as the time-limited format addressed difficulties associated with sustaining proximity to theraputic services for families in transition (Culhane et al., 2007). Given the innovative nature of this study, it should be noted that both the shelter provider and funders for this project addressed and overcame institutional skepticism to providing intensive services to sheltered children and families.

The mental and physical toll that the experience of homelessness has on caregivers (Arangua et al., 2005; Weinreb et al., 2006) undoubtably contributes to difficulties in the area of parenting (Koblinsky et al., 1997; McChesney, 1995; Fantuzzo & Perlman, 2007) as well as higher rates of parenting stress (Gorzka, 1999; Wu et al., 2018). These parenting difficulties are further exacerbated by the higher rates of social-emotional difficulties and EBP in their young children (e.g., Bussuk & Friedman, 2005). While emergency, transitional, or supportive housing programs for homeless families often provide parenting support services, the implementation of empirically supported parenting programs is quite rare (Gewirtz & Taylor, 2009). Our study shows that both time-limited CPP and PCIT significantly increased positive parenting verbalizations as well as improved/reduced overall parenting stress. Of note, while time-limited PCIT was effective in reducing the total and proportion of negative parenting verbalizations, time-limited CPP was only effective in reducing the proportion of negative parenting verbalizations (not the total) because parents’ overall verbalizations decreased at post. The fact that a 12-session time-limited version of CPP was moderately effective in changing some of these parenting outcomes is meaningful given that CPP was originally designed to be a year-long intervention (Lieberman et al., 2005). However, in terms of their relative effectiveness, the largest impact on mothers’ parenting (verbalizations and stress) came from those participating in time-limited PCIT which significantly outperformed time-limited CPP by a wide margin.

PCIT has been demonstrated to be effective in the treatment of children exposed to a variety of early childhood stressors including domestic violence (Borrego et al., 2008; Pearl, 2008); however, to our knowledge, the efficacy of PCIT has never been assessed within the context of homelessness. The current study adds to this literature by demonstrating that PCIT is effective in improving parenting and child outcomes in families currently experiencing homelessness. It also expands the literature on effective administration of PCIT with parents from a minority background as 78.5% of our mothers were Black/African-Americans and 27.1% were Hispanic/Latina. Finally, our study demonstrates the high transportability of PCIT, by showing that expensive resources (e.g., one-way mirrors, camera) are not necessary to intervention success, as the (masked for review) therapists provided live coaching in the same room as the family (e.g., play rooms at the shelter). These findings suggest that PCIT can be effectively and affordably administered within the context of homeless and other shelters and offers a promising avenue for addressing the pressing parenting needs of the most at-risk, in need, and underserved populations in our communities. These findings are particularly important in light of research indicating that homelessness is associated with increased parental frustration and negative parent-child interactions, decreased confidence in parenting, and difficulties with positive parent-child interactions (Koblinsky et al., 1997; Lee et al., 2010).

Consistent with our original hypothesis, PCIT outperformed CPP and was the only program to see statistically meaningful reductions in children’s EBP. Counter to our hypothesis, CPP did not outperform PCIT as it related to trauma symptom reductions. In fact, both parenting programs were equally effective in reducing children’s post-traumatic stress symptoms. The effects of these interventions were comparable to the effects of other well-known interventions of post-traumatic stress symptoms (Silverman et al., 2008); which is particularly noteworthy as PCIT was not originally designed as an intervention for childhood trauma. Although PCIT has been demonstrated to be effective in reducing child trauma symptoms (Pearl et al., 2012), to our knowledge, this is the first study to test this assertion within a randomized control trial and the first study to compare the effectiveness of PCIT with an intervention designed to address childhood trauma. The finding that PCIT was equally as effective as CPP in reducing child trauma symptoms provides further evidence that PCIT is not merely an intervention for EBP, but one that has a broader impact on early childhood psychopathology.

The exact mechanism by which PCIT addresses childhood trauma is as of yet unknown. However, targeting the parent-child relationship remains one of the core goals of trauma treatment in young children given their difficulty in verbally expressing past trauma (e.g., Lieberman et al., 2005). When considering the higher rates of EBP among children experiencing trauma (Cecil et al., 2017; Levendosky et al., 2002), PCIT’s dual focus on parent-child interactions and behavioral contingency management is particularly helpful. Certainly, the time-limited aspect of both interventions may also have influenced these results, in particular because the reduction in the number of sessions was greater for CPP than it was for PCIT (i.e., CPP was designed to be a yearlong intervention whereas PCIT is on average 20.5 sessions). Thus, it is unclear whether with a longer intervention period CPP may have had an impact on children’s EBP and/or performed better than PCIT in regards to addressing trauma symptoms. However, given the transient nature of sheltered families, the limited resources available, and the high demands within homeless shelters, time for intervention is at a premium making effective time-limited intervention protocols particularly valuable. As such, although the present study offers two viable intervention options for shelter staff aiming to address trauma symptoms in early childhood, PCIT may offer a more comprehensive intervention approach for addressing complex presentations with both EBP and trauma related concerns in this young age group.

In terms of our limitations, first, we cannot speak to the long-term maintenance of time-limited PCIT and CPP without follow-up data. The lack of follow-up was due to families exiting the shelter and limitations on the shelter’s resources. Second, it is important to acknowledge that we did not have a wait list control group given the service driven nature of this study. Thus, it is possible that some of the improvements seen in our parent and child outcomes were partially due to the infrastructure and supportive nature of the shelter. Third, the coding system (DPICS) used to measure mothers’ positive and negative verbalizations was originally developed for PCIT (Eyberg et al., 2013). Thus, perhaps it is not too surprising that mothers in PCIT outperformed those in CPP on those outcomes, although mothers in CPP also did improve significantly their positive verbalizations. Implementation of observational codes targeting more attachment related behaviors (e.g., general levels of warmth) that are less skills based is recommended for future work, although one recent study showed that these DPICS codes are moderately to highly correlated with some of the attachment related behavior codes (Blizzard et al., 2018). It will be important for future work to examine the efficacy of CPP and other programs targeting attachment in infancy among expecting mothers. Fourth, as with most community trials, it is important to note that 35% percent of our cases were led by a clinician cross-trained in both PCIT and CPP. While we covaried cross-training status in our analyses and implemented fidelity checklists as well as consistent supervision, it is always possible that some intervention contamination occurred. Finally, it is also important to acknowledge that most early intervention programs focus on the mother-child relationship/interaction (e.g., Eyberg et al., 2001; Sanders et al., 2000; Webster-Stratton et al., 2008), which is certainly the case when examining a women’s shelter of which only single mothers are served.

In terms of clinical implications, the current study shows the importance of offering early evidence-based assessments of the needs of sheltered children to detect and address their developmental and mental health needs, as well as the feasibility and effectiveness of embedding evidence-based parenting programs for early intervention within a homeless or domestic violence shelter. Overall, it shows that children and families within a shelter, whether due to homelessness, domestic violence or both, can benefit from CPP and PCIT programs in terms of not only reducing parenting stress but learning new parenting strategies that within a short period of time have significant benefits for children’s behavioral and emotional functioning. When considering that PCIT outperformed CPP on EBP and parenting outcomes (stress and maternal positive and negative verbalizations), time-limited PCIT may provide a more comprehensive and transdiagnostic service for sheltered families with preschool children. Future work should examine the extent to which changes in the parent-child relationship and/or stress mediate improvements seen in such a short time among sheltered families.

It is important to acknowledge that most agencies serving families experiencing homelessness are typically community-based with limited resources, high staff turnover, and little access to evidence-based programs (Gewirtz & August, 2008). Although providing families with basic needs remains the top priority at homeless shelters, there is a growing consensus on the importance of also addressing mental health needs (e.g., Bussuk & Friedman, 2005), particularly of vulnerable children. This study shows how a service driven, community-university partnership can play a large role in addressing the mental health needs of sheltered children and families with the potential to transform the trauma of homelessness into a window of opportunity.

## Data Availability

Data is available from the authors upon request.

## Acknowledgements

This project was made possible by the generous funding of: The Children’s Trust of Miami Dade County, Florida; Miami Dade County Homeless Trust; Miami Dade County and Lotus Endowment Fund, Inc.; Micky and Madeleine Arison Family Foundation; Carnival Foundation; Martin Z. Margulies; Angela Whitman and Family Foundation; and our community. We would like to thank the children and families of Lotus House for their participation and the dedicated team of Lotus House for making this community based, service driven research possible, with special mention to Shana Cox, LCSW, Psy.D., Gabrielle Contreras, LCSW, Stephanie Padro, LMFT, Muriel Ayala, LCSW, Nicole Carnero, LMHC, Nadly Moline, LCSW, Michaelle Sylveus, LMHC, Geneva Comeau, LMHC, Arleny Mirambeau, MSW, Marsha Trujillo, MFT, Ireysis Ramos Garcia, MSW, Franchesca Ali, MSW, LeShea Jenkins, MEd, Noelle Amador, BS, Leanett Reinoso, BA, and Melissa Claros-Erazo, BS.

## Footnotes

1 Rates of childhood trauma vary substantially based upon the specific traumatic events examined. Please see Saunders and Adams (2014) for a more detailed examination of the epidemiology of trauma in children and adolescents.

